# Validation of the German version of the Life-Space Assessment LSA-D

**DOI:** 10.1101/2021.01.13.21249715

**Authors:** Sandra A. Mümken, Paul Gellert, Malte Stollwerck, Julie L. O’Sullivan, Jörn Kiselev

## Abstract

**Objectives:** To develop a German version of the original University of Alabama at Birmingham (UAB) Study of Aging Life-Space Assessment (LSA-D) for measurement of community mobility in older adults within the past 4 weeks and to evaluate its psychometric properties for urban and rural populations of older adults.

**Design:** Cross-sectional validation study.

**Setting:** Two study centres in urban and rural German outpatient hospital settings.

**Participants:** In total N=83 community-dwelling older adults were recruited (n=40 from urban and n=43 from rural areas; mean age was 78.5 (SD=5.4); 49% male).

**Primary and secondary outcome measures:** The final version of the translated LSA-D was related with questions about activities and instrumental activities of daily living (ADL/iADL; primary hypothesis), Timed-Up&Go-Test (TUG), self-rated health, balance confidence and history of falls, use of transportation, and sociodemographic factors to obtain construct validity. Secondary outcome measures of health included handgrip strength, screening of cognitive function and comorbidities. To assess conduct construct validity, correlations between LSA-D and all health measures were examined for total sample, urban and rural subsamples using bivariate regression and multiple adjusted regression models. Posthoc analyses included different LSA-D scoring methods for each region. All parameters were estimated using non-parametric bootstrapping procedure.

**Results:** In the multiple adjusted model for the total sample, number of ADL/iADL limitations (β=-.26; 95%CI=-.42/-.08), TUG (β=-.37; 95%CI=-.68/-.14), living in shared living arrangements (β=.22; 95%CI=.01/.44) and history of falls in the past 6 months (β=-.22; 95%CI=-.41/-.05) showed significant associations with the LSA-D composite score, while living in urban area (β=-.19; 95%CI=-.42/.03) and male gender (β=.15; 95%CI=-.04/.35) were not significant.

**Conclusion:** The LSA-D is a valid tool for measuring life-space mobility in German community-dwelling older adults within the past four weeks in ambulant urban and rural settings.

**Trial registration number:** DRKS00019023

**Strengths and limitations of this study:**

- German validation of the original UAB Life-Space Assessment (LSA-D) for community dwelling older adults in urban and rural settings
- Using bootstrapped bivariate and multiple adjusted regression models to attain construct validity of the LSA-D
- Recruitment had to be stopped shortly before reaching the calculated sample size due to the decision to restrict in face-to-face research to contain the outbreak of the Covid-19 pandemic in March 2020

## Introduction

Mobility, defined as “the ability to move oneself (either independently or by using assistive devices or transportation) within environments that expand from one’s home to the neighbourhood and regions beyond” ^1^ encompasses general independence, opportunities for social activities and freedom to experience new sites. This broad concept of mobility goes beyond the narrow conception of mobility as performance in a single functional test without considering environmental barriers and social resources although their impact on mobility has been investigated.^2 3^ Therefore, the focus on single functional mobility tests can lead to misconceptions about actual mobility performance in everyday life and health practitioners may oversee possible consequences for social participation and mental health.^4^

To overcome these shortcomings of functional mobility assessments, recent studies of mobility and aging operationalize mobility as circled areas, so-called life-spaces, that spread from the centre of one’s own house and garden to the neighbourhood, the city lived in and beyond, with each life-space offering different opportunities for social involvement, recreational activities or access to medical care.^5 6^ The application of self-reported life-spaces to determine mobility of older adults was first established by May et al. in 1985 ^7^ and assessment of life-space mobility with standardized questionnaires was recently recommended for geriatric research.^8^ Several instruments for measuring life-space mobility in specific populations and settings exist, including assessments of life-space within one’s own residence for home-bound individuals ^9^ or residents in nursing homes and other institutions. ^10 11^

One of the most frequently applied instruments for measurement of mobility in older adults using the life-space concept is the validated Life-Space Assessment (LSA) by Baker, et al. ^12^ as part of the University of Alabama at Birmingham (UAB). Study of Aging. The LSA provides health professionals in geriatric settings with information on availability of environmental and social resources as an outcome of mobility assessment and gives them a more comprehensive picture of the patients’ needs.

The importance of the LSA for clinical practice has been shown in various studies. Kennedy et al. ^13^ for instance found that a decline in life-space over six months is associated with greater mortality in the following six months. Limitations in life-space mobility are associated with long-term mortality of older men ^14^, cognitive decline ^15^, risk of falls ^16^, frailty ^17^ and hospital admission in older adults with heart failure.^18^ Furthermore, the concept has already been established in outpatient physical therapy with various community-dwelling neurological orthopaedic and surgery patients.^19^ Additionally, psychological health factors like external control believes ^20^ and personal activity goals ^21^ influence life-space mobility. Therefore, the LSA can also supplement evaluation concepts in psychological research and treatment of older adults. The construct validity of the LSA was commonly tested by relating the LSA to activities an instrumental activities of daily living (ADL/iADL) but also self-rated health and fears of falling.^22-24^ Moreover, as Baker et al. ^12^ pointed out, there is a need to validate the LSA in urban and rural settings. As part of validity testing, the LSA has been translated into multiple languages such as Chinese ^25^, French ^24^, Spanish ^22^, Swedish ^26^ or Danish.^27^ To date, two modified German versions for assessment of life-space mobility in specific populations of older adults exist: One for those with mild cognitive impairment (LSA-CI) ^23^ that captures life-space of the past week and one for those in institutionalized settings where life-spaces are adapted to the living environment of care facilities (LSA-IS).^11^ However, a validated and intercultural adapted version of the original LSA that can be administered in the context of a more general geriatric setting in the overall population of community-dwelling older adults is still missing. Therefore, we conducted a validation study of a German version of the original LSA the (LSA-D) in urban and rural areas.

### Aims and Hypotheses

Our aim was to translate, apply and validate the LSA-D, a German version of the LSA from the University of Alabama (UAB) Study of Aging for the population of urban and rural community-dwelling older adults. In line with the original validation of the LSA we hypothesized a moderate association of the LSA-D composite score with limitations in ADL/iADL as primary hypothesis ^12^ Further, we assumed moderate associations with Timed-Up&Go-Test (TUG), self-rated health and history of falls. Finally, we expected the newly translated LSA-D to show these associations in both urban and rural populations of older adults.

## Methods

### Study design

A cross-sectional study design was used with two German hospital clinics as study centres. The first study centre was an ambulant geriatric rehabilitation facility of the Havelland clinics located in a small town (16,000 inhabitants) in Brandenburg, Germany. The second centre was based at the Charité – Universitätsmedizin Berlin within the Department of Anesthesiology and Operative Intensive Care. Approvement for the study was given by the local Ethics Committee of the Charité – Universitätsmedizin Berlin (EA2/124/19) and the study was prospectively registered at the German Clinical Trials Register (DRKS00019023). Sample size calculation was based on assumptions to find a moderate-to-strong association of beta/r=-.40 ^12^ between the LSA-D-composite score and ADL/iADL (i.e., primary hypothesis), functional mobility with TUG, self-rated health and balance confidence in all observed populations. For testing of the primary hypothesis, 92 participants or 46 subjects per setting (i.e., urban/rural) were required. This was based on the following assumptions: An effect size of Pearson’s correlation coefficient or standardized beta coefficient of r/beta=-0.40 (ρ=-0.40 in the population) was assumed in reference to the association between the LSA composite score (LS-C) and ADL/iADL found in the original validation study of LSA.^12^ The power calculation with GPower 3.1 for bivariate correlations (test family “exact”) ^28^ resulted in an estimated minimum sample size of n=46 participants per setting (urban/rural) and a critical r=-0.29 with a type I error rate of alpha=0.025 (test one-sided; corrected for multiple testing [setting urban/rural; alpha=0.05/2]) and a statistical power of 1-β=0.80. Recruitment took place from November 2019 to February 2020 and ended in March 2020 with a sample size of 82 due to restrictions of the then starting coronavirus pandemic. A post hoc sensitivity analysis suggests that we are still able to detect effects of r=-.30 and larger.

### Translation process

In accordance with the 2008 guidelines of the World Health Organization ^29^, forward translation into German language was separately conducted by two researchers who formulated two German versions that were discussed and then merged into one German pre-version of the LSA-D. The pre-version was given to two native English speakers for back translation. Again, both versions of the back translation were discussed by the two native speakers and a concerted version of the back-translation was produced. Differences between the original LSA and the concerted back-translation were discussed and reviewed with the original author of the LSA to redefine a pre-final version of the LSA-D that was pre-tested for understandability using cognitive interview technique among 4 older adults of the Charité – Universitätsmedizin Berlin to create the final LSA-D version.^30^

### Participants and Recruitment

The 83 participants were divided into two groups based on the size of their place of residence. Participants from villages (< 5,000 inhabitants) and small towns (up to 40,000 inhabitants) were classified as living in rural areas. In contrast, participants who lived in the city of Berlin (3.8 million inhabitants) with its metropolitan infrastructure and services were classified as urban population. Inclusion criteria were defined as: age of 70 years and older; ability to read and understand the questionnaire and give written informed consent. Exclusion criteria were incidences that limited mobility within the past four weeks, severe cognitive limitations or mental conditions, need of acute care and insufficient understanding of the German language. In total, 126 persons were screened for eligibility of which 28 did not fulfil the inclusion criteria and 15 were unwilling to participate. In both study centres, participants were recruited during normal health care routine by trained study staff. All participants received verbal and written information on the study and were given time to consider participation before giving written consent.

## Measures

### Sociodemographic measures

Demographic variables (i.e., age, gender, height, weight, status of shared living-arrangements) use of public transportation and driving-status were assessed with a standardized questionnaire.

### Primary outcome measures

Selection of other primary and secondary variables for determining construct validity was based on the original validation study of the UAB and other LSA validation studies from different countries.^12 22-24^

Life-space was evaluated with the translated German Version of the UAB Life-Space Assessment. The LSA consists of a questionnaire on five different life-spaces capturing six possible levels of life-space (0. Mobility within the bedroom, 1. rooms inside the home besides the bedroom, 2. area outside the house, 3. neighbourhood, 4. town or city lived in, outside of town or city lived in). For each level, participants were asked a) if they went to this level in the past four weeks, b) if so, how often, c) if they needed assistive devices or special equipment to reach that level and d) if they needed personal help to reach that level. ^12^ Different scoring methods can be used with the LSA either indicating the maximum attained life-space level (LS-M), life-space that can be reached independently without any further support (LS-I), reachable life-space with possible use of equipment but without personal help (LS-E), dichotomized life-space (LS-D) that classifies a person’s mobility into the ability to travel beyond the borders of their self-perceived neighbourhood and the composite score (LS-C) that summarizes the attained LS-level, needed equipment or personal support and frequency of visits. The LS-C score is calculated with a defined algorithm and ranges from 0 to 120 points with higher scores indicating better mobility. As the LS-C score has shown a good sensitivity regarding change over time, it is frequently applied in longitudinal studies. ^31 32^ In cross-sectional studies, LS-I and LS-D are additional scores for describing actual mobility and associations with other health factors. ^12^

Limitations in ADL and iADL were investigated using questions from the “Survey of Aging and Retirement in Europe” (SHARE).^33^ Participants were asked whether they had difficulties due to physical, emotional or cognitive problems to perform 15 activities like dressing, toileting, gardening, using a map or making a telephone call. Binary response options for each activity were yes or no. Subsequently, a sum score of limitations in ADL/iADL activities was calculated ranging from 0 to 15. Higher scores indicate more functional impairments.

The TUG is one of the most frequently used measures of balance and functional mobility in older adults and is a recommended tool for geriatric assessment.^34^ During performance of the TUG, time (in seconds) is taken for rising up from a standardized chair, walking three metres, turning around, walking back and siting down again in a comfortable self-selected speed. ^35^ Higher TUG times are associated with stronger mobility and ADL restrictions.^36 37^

The EQ-VAS from the EQ-5D-5L version was used to record overall self-rated health of the day on a vertical visual analogue scale ranging from 0 points for the worst imaginable health to 100 points for the best conceivable health.^38^ To measure balance confidence, we used the ABC-6-Scale that was translated into German and validated by Schott et al.^39^ Participants were accounted to have a history of falls if they had fallen at least one time in the past six months defining a fall as ‘‘unintentionally coming to rest on ground, floor, or other lower level; excludes coming to rest against furniture, wall, or other structure.”^40^.

### Secondary outcome measures of health

Hand grip strength was measured as maximum of three contractions with a hydraulic handheld dynamometer (Sahean SH5001; Changwon, South Korea) in the dominant hand and standardized sitting position.^41^ We administered the Charlson comorbidity index (CCI) as a method to categorize comorbidities as each of its 17 items is weighted based on the adjusted risk for mortality and resource use, leading to a score of 0-41 points and scores of >5 indicate a higher mortality risk.^42^ Cognitive status was assessed with the Mini-Cog screening tool where a score ranging from 0-5 can be achieved and a score of 0-2 is seen as an indicator for further investigation of cognitive status.^43^

### Statistical analysis

Means (M) and standard deviations (SD) were reported descriptively for continuous demographic variables (i.e., age, height, weight) and health measures (i.e., limitations in ADL/iADL, time in seconds needed to complete TUG, self-rated health and balance confidence). Gender, status of shared living arrangements, use of different transportation modes and history of falls were reported for the total and each subsample as absolute frequencies and percentage of participants. Distribution of the data was skewed therefore we used the non-parametric, bias corrected and accelerated (BCA) bootstrap method with 10,000 resamples and fixed random seeds that resamples the collected data with replacement to derive robust results.^44^ With the BCA bootstrap method, coefficients and confidence intervals can be estimated with good statistical power even if sample sizes are small and distribution of data is unknown or not normal. For investigating differences between urban and rural participants, the Welch Test was performed as it has been recommended as a standard test for small samples.^45^ To determine construct validity of the LSA-D, BCA bootstrap method and standardized z-scores (i.e., that can be interpreted like beta coefficients) of the included binary and continuous variables (i.e., age, male gender, rural or urban residence, status of shared living arrangements, sum score of limitations with ADL/iADL activities, functional mobility with TUG, self-rated health, balance confidence and history of falls) were used for bivariate and multivariate regression analysis. Scores of the ABC-6 scale had to be excluded from multivariate regression because they revealed a correlation of r=-.72 with TUG scores. To avoid multicollinearity, it was decided to include only the TUG score due to its importance as a physical measurement of functional mobility for assessing construct validity. All analyses were run using SPSS version 25. Microsoft Excel 2016 was used to create the figure.

## Results

### Sample characteristics

For the total sample (N=83), mean age was 78.5 (SD=5.4) years and about half of the sample (n=41; 49.4%) were male. 47 participants (56.6%) lived together with others in a shared living arrangement. In the past four weeks, 39 participants (47.0%) drove a car by themselves, 18 participants (21.7%) rode a bicycle and 34 participants (41.0%) used walking aids. On average, participants had a TUG of M=13.90 (SD=9.20) seconds. Score of limitations in ADL/iADL was moderate with M=7.8 (SD=6.2) and mean score of self-rated health was M=64.7 (SD=21.3).

When comparing urban with rural participants, those living in urban areas had significantly more ADL/iADL limitations, t(74.51)=-2.34; p=.022, and comorbidities, t(57.27)=-2.44; p=.018. Rural participants were significantly older, t(81)=2.43; p=.017, needed more time to complete the TUG, t(70.65)=3.33; p=.001, had less balance confidence, t(80.11)=-2.84; p=.006, were more often assigned to a care level, t(69.62)=4.53; p<.001, and had lower self-rated health, t(81)=-2.45; p=.016. Concerning the utilization of means of transportation, the percentage of participants who drove a car or a bicycle for independent mobility within the last 4 weeks did not differ significantly across regions. Characteristics of participants in total and separately for each region are presented in table 1.

**Table 1:** Participant Characteristics

|  | total (N=83) |  | urban (n=40) |  | rural (n=43) |  |
| --- | --- | --- | --- | --- | --- | --- |
| Variable | *N* | % | *N* | % | *N* | % |
| Gender (male) | 41 | 49.4 | 23 | 57.5 | 18 | 41.9 |
| Status of shared living arrangements | 47 | 56.6 | 19 | 47.5 | 28 | 65.1 |
| Drove a car in past 4 weeks | 39 | 47.0 | 18 | 45.0 | 21 | 48.8 |
| Rode a bicycle in past 4 weeks | 18 | 21.7 | 9 | 22.5 | 9 | 20.9 |
| Used walking aid in past 4 weeks | 34 | 41.0 | 11 | 27.5 | 23 | 53.5 |
| History of falls past 6 months (>1) | 22 | 26.5 | 9 | 22.5 | 13 | 30.2 |
|  | *M* | *SD* | *M* | *SD* | *M* | *SD* |
| Age (years) | 78.5 | 5.4 | 77.1 | 5.2 | 79.8 | 5.2 |
| Height (cm) | 168.7 (3) | 10.5 | 170.8 | 11.2 | 166.5 (3) | 9.3 |
| Weight (kg) | 79.0 (3) | 18.8 | 79.7 | 19.8 | 78.3 (3) | 17.8 |
| Body Mass Index | 27.6 (3) | 5.4 | 27.1 | 5.8 | 28.1 (3) | 5.1 |
| Charlson Comorbidity Index (RANGE) | 3.2 | 3.6 | 4.2 | 4.5 | 2.3 | 2.3 |
| Hand grip strength (kg) | 25.8 (2) | 11.6 | 27.7 (1) | 11.8 | 24.0 (1) | 11.4 |
| Mini-Cog™-Score (0-5) | 3.8 | 1.5 | 3.8 | 1.6 | 3.8 | 1.4 |
| LSA-D composite score (0-120) | 60.8 | 24.3 | 58.0 | 21.7 | 63.3 | 26.5 |
| ADL/iADL (number of limitations) | 7.8 | 6.2 | 9.4 | 6.7 | 6.2 | 5.4 |
| TUG (s) | 13.9 (5) | 9.2 | 10.5 (3) | 6.8 | 16.9 (2) | 10.1 |
| Self-rated health (0-100) | 64.8 | 21.3 | 70.5 | 19.3 | 59.4 | 21.8 |
| Balance confidence (0-100) | 57.8 | 32.7 | 67.8 | 28.4 | 48.4 | 34.0 |

Note: Live-Space-Assessment-Deutsch (LSA-D); Activities of Daily Living (ADL); Instrumented Activities of Daily Living (iADL); Timed-Up &Go-Test (TUG); Numbers in ( ) report number of missing values

### Descriptive statistics of the LSA-D

Life-space-level 5, as maximum life-space (LS-M), was reached by 60.2% of the total sample. 32.5% of all participants had an independent life-space level (LS-I) of 5 while the remaining needed either equipment or personal help. For the urban subsample, 40.0% of the participants reached life-space level 5 as LS-M and 27.5% of urban participants reached life-space level 5 independently without any support (LS-I). In contrast, 79.1% of rural participants achieved LS-level 5 as LS-M and 37.2% did this independently without any support (LS-I). Figure 1 illustrates the different life-space measures among the total sample and urban/rural subsample. No significant differences between urban and rural participants were observed in LS-C: (t(81)=1.00; p=.323), LS-E: (t(81)=0.57; p=.571), and LS-D: (t(80.99)=-1.95; p=.054). Rural participants had a significantly higher LS-M: (t(64.60)=3.83; p<.001), and LS-I: (t(77)=-2.00; p=.049).

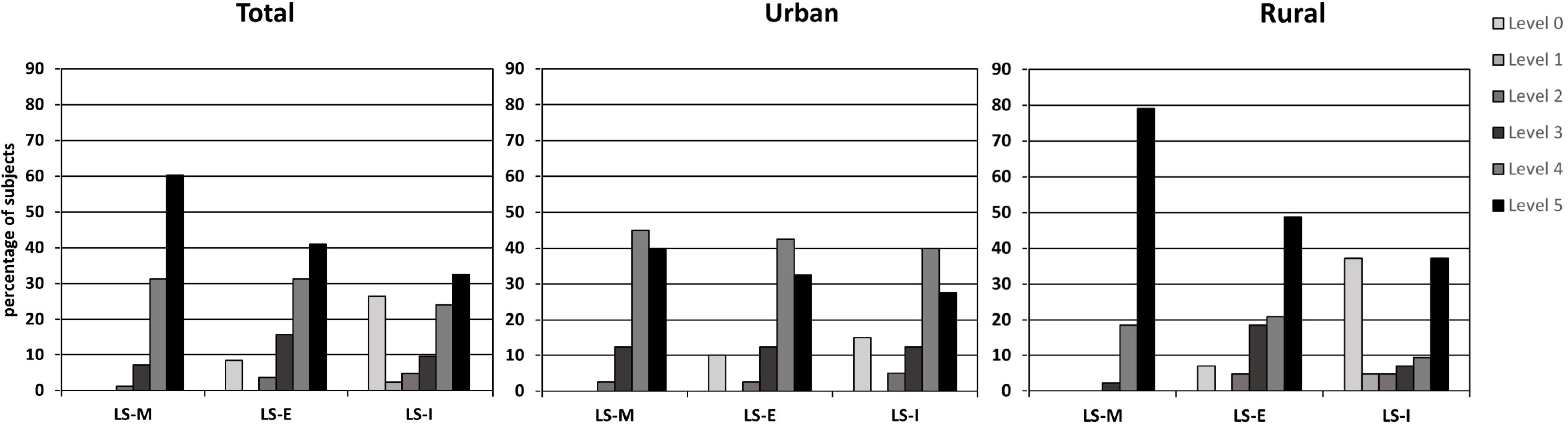

### Construct validity

For the total sample, associations from the bivariate regression analyses between the LSA-D composite score, demographic variables, functional mobility and other health measures were significant for age (β=-.24; 95%CI=-.44/-.07; p=.016), status of shared living arrangements (β=.22; 95%CI=.01/.43; p=.040) ADL/iADL (β=-.23; 95%CI=-.43/-.01; p=.034), TUG (β=-.47; 95%CI=-.66/-.34; p<.001), self-rated health (β=.40; 95%Cl=.19/.61; p<.001), history of falls (β=-.35; 95%CI=-.54/-.15; p<.001). Male gender (β=-.09; 95%CI=-.31/.13; p=.407) and urban residence (β=-.11; 95%CI=-.33/.10; p=.314) were not significant for the total sample. In the adjusted model, age, male gender, urban residence, status of shared living arrangements, number of limitations in ADL/iADL, TUG, self-rated health and history of falls were included into the equation in one step for the total sample. The result revealed significant associations for living status in shared living arrangements (β=.22; 95%CI=.01/.44; p=.045), limitations in ADL/iADL activities (β=-.26; 95%CI=-.42/-.08; p=.008), functional mobility measured with the TUG (β=-.37; 95%CI=-.68/-.14; p=.008) and history of falls (β=-.22; 95%CI=-.41/-.05; p=.018). No significant associations were found for male gender (β=.15; 95%CI=-.04/.35; p=.135) and urban residence (β=-.19; 95%CI=-.42/.03; p=.090), which corresponds with the bivariate model. However, in contrast to bivariate models, influence of age (β=-.08; 95%CI=-.32/.12; p=.509) and self-rated health (β=.24; 95%CI=.02/.47, p=.058) were not significant in the multivariate model. Results of bivariate and adjusted multivariate regression models are shown in table 2.

**Table 2:** Unadjusted and adjusted associations of sociodemographic and health factors with the LSA-D composite score (n=83)

| Variable | *Bivariate unadjusted models* |  |  | *Adjusted model* |  |  |
| --- | --- | --- | --- | --- | --- | --- |
|  | *Beta* | *LL CI / UL CI* | *p* | *Beta* | *LL CI / UL CI* | *p* |
| Age | **-.24** | **-.44/-.07** | **.016** | -.08 | -.32/.12 | .509 |
| Gender (male) | -.09 | -.31/.13 | .407 | .15 | -.04/.35 | .135 |
| Living in shared living arrangements | **.22** | **.01/.43** | **.040** | **.22** | **.01/.44** | **.045** |
| Lives in urban area | -.11 | -.33/.10 | .314 | -.19 | -.42/.03 | .090 |
| ADL/iADL-Score | **-.23** | **-.43/-.01** | **.034** | **-.26** | **-.42/-.08** | **.008** |
| TUG | **-.47** | **-.66/-.34** | **< .001** | **-.37** | **-.68/-.14** | **.008** |
| Self-rated health | **.40** | **.19/.61** | **< .001** | .24 | .02/.47 | .058 |
| Balance confidence | **.50** | **.33/.67** | **< .001** | - | - | - |
| History of falls in past 6 months | **-.35** | **-.54/-.15** | **< .001** | **-.22** | **-.41/-.05** | **.018** |

Notes: Timed-Up&Go-Test (TUG); Activities of Daily Living (ADL); Instrumental Activities of Daily Living (iADL); p < .05 highlighted in bold

Separate bivariate regression analyses for the urban and rural region demonstrated comparable results in the urban and rural subsample for TUG urban: (β=-.48; 95%CI=-1.14/-.32; p=.008) and rural: (β=-.60; 95%CI=-.95/-.40; p<.001), self-rated health urban: (β=.51; 95%CI=.29/.90; p=.001) and rural: (β=.43; 95%CI=.12/.77; p=.010), balance confidence urban: (β=.67; 95%CI=.38/.93; p=<.001) and rural: (β=.54; 95%CI=.29/.81; p=.001) and history of falls urban: (β=-.31; 95%CI=-.59/-.03; p=.030) and rural: (β=-.41; 95%CI=-.67/-.11; p=.009). Age was significant for those living in the urban region (β=-.31; 95%CL=-.53/-.09; p=.011), but not for the rural sample (β=-.28; 95%CI=- .68/.03; p=.147). All other demographic variables and health measures showed no significant associations in both groups. Results are presented in table 3.

**Table 3:** Unadjusted associations of sociodemographic and health factors with LSA-D composite score for participants in urban (n=40) and rural areas (n=43)

| Variable | *Urban* |  |  | *Rural* |  |  |
| --- | --- | --- | --- | --- | --- | --- |
|  | *Beta* | *LL CI / UL CI* | *p* | *Beta* | *LL CI / UL CI* | *P* |
| Age | **-.31** | **-.53/-.09** | **.011** | -.28 | -.68/.03 | .147 |
| Gender (male) | -.20 | -.43/.06 | .151 | -.04 | -.38/.29 | .826 |
| Living in shared living arrangements | .18 | -.11/.45 | .231 | .24 | -.10/.54 | .150 |
| ADL/iADL-Score | -.16 | -.39/.11 | .209 | -.28 | -.71/.11 | .208 |
| TUG | **-.48** | **-1.14/-.32** | **.008** | **-.60** | **-.95/-.40** | **< .001** |
| Self-rated health | **.51** | **.29/.90** | **.001** | **.43** | **.12/.77** | **.010** |
| Balance confidence | **.67** | **.38/.93** | **< .001** | **.54** | **.29/.81** | **.001** |
| History of falls in past 6 month | **-.31** | **-.59/-.03** | **.030** | **-.41** | **-.67/-.11** | **.009** |

Notes: Timed-Up&Go-Test (TUG); Activities of Daily Living (ADL); Instrumental Activities of Daily Living (iADL); p < .05 highlighted in bold

Post-hoc calculations of the adjusted model for each sample separately showed a significant regression coefficient in the urban sample for the score of ADL/iADL limitations (β=-.23; 95%CI=-.41/-.10; p=.035) while coefficients for all other variables were not significant. For the rural population, results for status of shared living arrangements (β=.39; 95%CI=.02/.75; p=.039) and history of falls (β=-.42; 95%CI=-.80/-.08; p=.04) showed significance while other variables did not.

## Discussion

We translated and validated a German version of the LSA in urban and rural community-dwelling older adults. In line with the original validation of the LSA ^12^, moderate associations of the LSA-D composite score with status of living in shared living arrangements, limitations in ADL/iADL as well as with physical performance assessed with the TUG and history of falls were found in the standardized adjusted model. The standardized adjusted association of limitations in ADL/iADL with the LSA-D composite score revealed in our study is in line with findings of Baker et al. ^12^ and Curcio ^22^, albeit lower than expected. We found stronger moderate adjusted associations for functional mobility measured with the TUG. These results correspond with findings by Ullrich et al. who reported a moderate Pearson correlation with the TUG when validating the modified LSA-CI to capture life-space of older adults with mild cognitive impairment during the past week.^46^ Previous validation studies have tested their version of the LSA composite score against against the Short Physical Performance Battery as a physical assessment of functional mobility and found moderate to strong association.^12 26^ Furthermore, our results revealed a moderate significant association for self-rated health with the LSA-D composite score in bivariate regression, which is in accordance with the original LSA validation study.^12^ However, this association did not remain significant in the adjusted model. Unfortunately, balance confidence as an additional subjective health measure that showed moderate significant bivariate associations could not be included in the adjusted model due to multicollinearity.

Our adjusted model confirms the importance of social resources as they can be seen in living together with others in shared living arrangements and functional mobility represented by the significant negative associations with limitations in ADL/iADL activities, time to complete the TUG and a positive history of falls. The importance of social resources was strengthened in post-hoc analyses, where we calculated the adjusted model for each regional subsample separately. Results showed that living in shared living arrangements and history of falls were significantly associated with life-space mobility in rural areas. This indicates that a nearby social network may play an even more important role in rural areas. In this regard, Kusipar et al. ^5^ also found evidence for the importance of social support on LS-mobility in the Canadian Longitudinal Study of Aging. Taken together, our findings demonstrate robust evidence for a good construct validity of the LSA-D.

To test our hypothesis that the LSA-D is applicable in both urban and rural living environments, we calculated separate bivariate regressions for each subsample. The associations were similarly strong for functional mobility, self-rated health, balance confidence and history of falls. Although a significant association with age was only found for the urban population and limitations in ADL/iADL were not significant in either subsample, our findings generally correspond across both subsamples. This supports our notion that the LSA-D can be used for measurement of life-space mobility during the past four weeks in community dwelling older adults living in both urban and rural areas.

To further determine construct validity of the LSA-D, we investigated the different scoring methods in the total and both subsamples separately. The LS-M differed between urban and rural participants, with those from rural areas reporting higher LS-M than those living in urban surroundings. Older adults living in rural areas might be more dependent on leaving their village or town in order to gain access to health care services or to run routine errands due to the limited infrastructure often found in rural areas. Our results suggest that the LSA-D is a useful tool for capturing specific characteristics of urban and rural living environments. No group differences were found concerning the LSA-D composite score and the LS-D soring method. This demonstrates the ability of the LSA-D composite score and the dichotomized LS-D score to remain stable and applicable outcome measures in urban and rural living environments.

### Strengths and limitations

A main strength of our study is that we tested psychometric properties of the LSA-D among urban and rural community-dwelling older adults. As mobility patterns may vary across living areas, assessments of mobility need to be valid for people living in small villages and large cities as well. Our findings revealed an urban-rural difference in maximal LS and thus demonstrate that the LSA-D can detect disparities in individual mobility patterns that are related to the surrounding living area. These differences must be considered when health care practitioners or researchers address specific questions about independence, social support and functional mobility in different regions. Maximal LS measured with the LS-M score is likely to vary between urban and rural areas and thus may reflect availability of different environmental resources and social support. Another strength is that we applied advanced statistical methods including non-parametric bootstrapping procedures and multivariate regression analysis to account for confounding variables and to estimate the independent association of each of the variables with LSA-D composite-score. However, there are some limitations that need to be considered. First, due to the beginning of the Covid-19 pandemic, we did not reach the planned sample size. However, post-hoc sensitivity analyses revealed a sufficient statistical power. Second, although statistical power was sufficient and the focus was on community-dwelling older adults, our sample size was rather small and non-representative. Future studies should replicate our findings in a representative sample, including subgroups of older adults with mild cognitive impairment, care dependency or living in nursing care arrangements. Moreover, future studies should consider the overlap between LS scoring methods of the LSA-D, social constructs and objectively derived GPS data. This multimodal approach can lead to a better understanding of complex mobility patterns in older adults and associated factors.

## Conclusion

In conclusion, the LSA-D has shown good construct validity and can be used in the general population of community-dwelling older adults in urban and rural living environments. The use of LSA-D is recommended for geriatric health care practitioners of different disciplines to assess mobility in the context of social participation and health service utilization.

## Supporting information

LSA-D Questionaire

## Data Availability

Data will be made available on reasonable, methodologically sound request as soon as possible to achieve the aims of the approved proposal.
Available data includes individual participant data that underlies the results reported in this article after its deidentification, data dictionary and the analytical code. 
Data will be available at least 3 months after, and ending 5 years following article publication. Proposals should be directed at

## Acknowledgments

We sincerely thank Patricia Sawyer for her feedback on the pre-final, back translated English LSA-D version. Also, we would like to thank Monica Schanzer and Rudolf Mörgeli for the back translation of the LSA-D, all participants and all members of the study staff that are not listed as authors.

## Footnotes

### Contributors

JK developed the first draft of the study. JK, SAM and PG planned and conducted the final studyprotocol. JK and SAM translated LSA into German, compiled the final version of the LSA-D, and contributed in recruiting and testing participants. SAM drafted the manuscript and conducted the major analyses. MS drafted tables and the figure. PG established the statistical analysis plan and performed the sample size and power calculation. SAM, and MS assisted with data analyses and interpretation of the results. JLOS gave advice and commented on the manuscript. All authors critically reviewed the manuscript and approved the final version.

### Patient consent for publication

Non declared.

### Ethical approval

Ethics approval for the study was provided by the Ethics Committee of Charité – Universitätsmedizin Berlin (Rec. Reference EA2/124/19).

### Data availability statement

Data will be made available on reasonable, methodologically sound request as soon as possible to achieve the aims of the approved proposal.

Available data includes individual participant data that underlies the results reported in this article after its deidentification, data dictionary and the analytical code.

Data will be available at least 3 months after, and ending 5 years following article publication. Proposals should be directed at

## References

1. Webber SC, Porter MM, Menec VH. Mobility in older adults: a comprehensive framework. Gerontologist 2010;50(4):443–50. doi: 10.1093/geront/gnq013 [published Online First: 2010/02/11]

2. Rantakokko M, Wilkie R. The role of environmental factors for the onset of restricted mobility outside the home among older adults with osteoarthritis: a prospective cohort study. BMJ Open 2017;7(6):e012826. doi: 10.1136/bmjopen-2016-012826

3. Spaltenstein J, Bula C, Santos-Eggimann B, et al. Factors associated with going outdoors frequently: a cross-sectional study among Swiss community-dwelling older adults. BMJ Open 2020;10(8):e034248. doi: 10.1136/bmjopen-2019-034248

4. Giannouli E, Bock O, Mellone S, et al. Mobility in Old Age: Capacity Is Not Performance. BioMed research international 2016;2016:3261567–67. doi: 10.1155/2016/3261567 [published Online First: 2016/02/29]

5. Kuspinar A, Verschoor CP, Beauchamp MK, et al. Modifiable factors related to life-space mobility in community-dwelling older adults: results from the Canadian Longitudinal Study on Aging. BMC Geriatr 2020;20(1):35. doi: 10.1186/s12877-020-1431-5 [published Online First: 2020/02/02]

6. Portegijs E, Keskinen KE, Tsai L-T, et al. Physical Limitations, Walkability, Perceived Environmental Facilitators and Physical Activity of Older Adults in Finland. International Journal of Environmental Research and Public Health 2017;14(3):333. doi: 10.3390/ijerph14030333

7. May D, Nayak US, Isaacs B. The life-space diary: a measure of mobility in old people at home. Int Rehabil Med 1985;7(4):182-6. [published Online First: 1985/01/01]

8. Taylor JK, Buchan IE, van der Veer SN. Assessing life-space mobility for a more holistic view on wellbeing in geriatric research and clinical practice. Aging Clinical and Experimental Research 2019;31(4):439–45. doi: 10.1007/s40520-018-0999-5

9. Hashidate H, Shimada H, Shiomi T, et al. Measuring indoor life-space mobility at home in older adults with difficulty to perform outdoor activities. J Geriatr Phys Ther 2013;36(3):109–14. doi: 10.1519/JPT.0b013e31826e7d33 [published Online First: 2012/09/15]

10. Tinetti ME, Ginter SF. The nursing home life-space diameter. A measure of extent and frequency of mobility among nursing home residents. J Am Geriatr Soc 1990;38(12):1311–5. doi: 10.1111/j.1532-5415.1990.tb03453.x [published Online First: 1990/12/01]

11. Hauer K, Ullrich P, Heldmann P, et al. Validation of the interview-based life-space assessment in institutionalized settings (LSA-IS) for older persons with and without cognitive impairment. BMC Geriatrics 2020;20(1):534. doi: 10.1186/s12877-020-01927-8

12. Baker PS, Bodner EV, Allman RM. Measuring life-space mobility in community-dwelling older adults. J Am Geriatr Soc 2003;51(11):1610-4. [published Online First: 2003/12/23]

13. Kennedy RE, Sawyer P, Williams CP, et al. Life-Space Mobility Change Predicts 6-Month Mortality. J Am Geriatr Soc 2017;65(4):833–38. doi: 10.1111/jgs.14738 [published Online First: 2017/02/06]

14. Mackey DC, Cauley JA, Barrett-Connor E, et al. Life-space mobility and mortality in older men: a prospective cohort study. J Am Geriatr Soc 2014;62(7):1288–96. doi: 10.1111/jgs.12892 [published Online First: 2014/06/18]

15. Silberschmidt S, Kumar A, Raji MM, et al. Life-Space Mobility and Cognitive Decline Among Mexican Americans Aged 75 Years and Older. J Am Geriatr Soc 2017;65(7):1514–20. doi: 10.1111/jgs.14829 [published Online First: 2017/03/10]

16. Lo AX, Rundle AG, Buys D, et al. Neighborhood Disadvantage and Life-Space Mobility Are Associated with Incident Falls in Community-Dwelling Older Adults. J Am Geriatr Soc 2016;64(11):2218–25. doi: 10.1111/jgs.14353 [published Online First: 2016/11/22]

17. Portegijs E, Rantakokko M, Viljanen A, et al. Is frailty associated with life-space mobility and perceived autonomy in participation outdoors? A longitudinal study. Age Ageing 2016;45(4):550–3. doi: 10.1093/ageing/afw072 [published Online First: 2016/04/30]

18. Lo AX, Flood KL, Kennedy RE, et al. The Association Between Life-Space and Health Care Utilization in Older Adults with Heart Failure. J Gerontol A Biol Sci Med Sci 2015;70(11):1442–7. doi: 10.1093/gerona/glv076 [published Online First: 2015/07/30]

19. McCrone A, Smith A, Hooper J, et al. The Life-Space Assessment Measure of Functional Mobility Has Utility in Community-Based Physical Therapist Practice in the United Kingdom. Physical Therapy 2019;99(12):1719–31. doi: 10.1093/ptj/pzz131

20. Sartori AC, Wadley VG, Clay OJ, et al. The relationship between cognitive function and life space: the potential role of personal control beliefs. Psychol Aging 2012;27(2):364–74. doi: 10.1037/a0025212 [published Online First: 2011/08/31]

21. Saajanaho M, Rantakokko M, Portegijs E, et al. Personal goals and changes in life-space mobility among older people. Prev Med 2015;81:163–7. doi: 10.1016/j.ypmed.2015.08.015 [published Online First: 2015/09/09]

22. Curcio CL, Alvarado BE, Gomez F, et al. Life-Space Assessment scale to assess mobility: validation in Latin American older women and men. Aging Clin Exp Res 2013;25(5):553–60. doi: 10.1007/s40520-013-0121-y [published Online First: 2013/08/21]

23. Ullrich P, Werner C, Bongartz M, et al. Validation of a Modified Life-Space Assessment in Multimorbid Older Persons With Cognitive Impairment. Gerontologist 2019;59(2):e66–e75. doi: 10.1093/geront/gnx214 [published Online First: 2018/02/03]

24. Auger C, Demers L, Gelinas I, et al. Development of a French-Canadian version of the Life-Space Assessment (LSA-F): content validity, reliability and applicability for power mobility device users. Disabil Rehabil Assist Technol 2009;4(1):31–41. doi: 10.1080/17483100802543064 [published Online First: 2009/01/28]

25. Tseng YC, Gau BS, Lou MF. Validation of the Chinese version of the Life-Space Assessment in community-dwelling older adults. Geriatr Nurs 2020;41(4):381–86. doi: 10.1016/j.gerinurse.2019.11.014 [published Online First: 2019/12/11]

26. Fristedt S, Kammerlind AS, Bravell ME, et al. Concurrent validity of the Swedish version of the life-space assessment questionnaire. BMC Geriatr 2016;16(1):181. doi: 10.1186/s12877-016-0357-4 [published Online First: 2016/11/09]

27. Pedersen MM, Kjaer-Sorensen P, Midtgaard J, et al. A Danish version of the life-space assessment (LSA-DK) - translation, content validity and cultural adaptation using cognitive interviewing in older mobility limited adults. BMC Geriatr 2019;19(1):312. doi: 10.1186/s12877-019-1347-0 [published Online First: 2019/11/16]

28. Faul F, Erdfelder E, Buchner A, et al. Statistical power analyses using G*Power 3.1: tests for correlation and regression analyses. Behav Res Methods 2009;41(4):1149–60. doi: 10.3758/brm.41.4.1149 [published Online First: 2009/11/10]

29. WHO. Process of translation and adaption of instruments https://www.who.int/substance_abuse/research_tools/translation/en/2008 [accessed 2020_12_17 2020.

30. Forsyth BHK, Martha Stapelton; Lawrence, Deidre; Levine, Kerry; Willis Gorgon B. Methods for Translating Survey Questionaires. American Association for Public Opinion Research 2006

31. Ritchie CS, Locher JL, Roth DL, et al. Unintentional weight loss predicts decline in activities of daily living function and life-space mobility over 4 years among community-dwelling older adults. J Gerontol A Biol Sci Med Sci 2008;63(1):67–75. doi: 10.1093/gerona/63.1.67 [published Online First: 2008/02/05]

32. Sheppard KD, Sawyer P, Ritchie CS, et al. Life-space mobility predicts nursing home admission over 6 years. J Aging Health 2013;25(6):907–20. doi: 10.1177/0898264313497507 [published Online First: 2013/08/24]

33. Borsch-Supan A, Brandt M, Hunkler C, et al. Data Resource Profile: the Survey of Health, Ageing and Retirement in Europe (SHARE). Int J Epidemiol 2013;42(4):992–1001. doi: 10.1093/ije/dyt088 [published Online First: 2013/06/20]

34. Turner G, Clegg A. Best practice guidelines for the management of frailty: a British Geriatrics Society, Age UK and Royal College of General Practitioners report. Age and Ageing 2014;43(6):744–47. doi: 10.1093/ageing/afu138

35. Podsiadlo D, Richardson S. The timed “Up & Go”: a test of basic functional mobility for frail elderly persons. J Am Geriatr Soc 1991;39(2):142–8. doi: 10.1111/j.1532-5415.1991.tb01616.x [published Online First: 1991/02/01]

36. Lin MR, Hwang HF, Hu MH, et al. Psychometric comparisons of the timed up and go, one- leg stand, functional reach, and Tinetti balance measures in community-dwelling older people. J Am Geriatr Soc 2004;52(8):1343–8. doi: 10.1111/j.1532-5415.2004.52366.x [published Online First: 2004/07/24]

37. Donoghue OA, Savva GM, Cronin H, et al. Using timed up and go and usual gait speed to predict incident disability in daily activities among community-dwelling adults aged 65 and older. Arch Phys Med Rehabil 2014;95(10):1954–61. doi: 10.1016/j.apmr.2014.06.008 [published Online First: 2014/07/01]

38. Ludwig K, Graf von der Schulenburg JM, Greiner W. German Value Set for the EQ-5D-5L. PharmacoEconomics 2018;36(6):663–74. doi: 10.1007/s40273-018-0615-8

39. Schott N. Reliability and validity of the German short version of the Activities specific Balance Confidence (ABC-D6) scale in older adults. Arch Gerontol Geriatr 2014;59(2):272–9. doi: 10.1016/j.archger.2014.05.003 [published Online First: 2014/06/26]

40. Buchner DM, Hornbrook MC, Kutner NG, et al. Development of the common data base for the FICSIT trials. J Am Geriatr Soc 1993;41(3):297–308. doi: 10.1111/j.1532-5415.1993.tb06708.x [published Online First: 1993/03/01]

41. Sousa-Santos AR, Amaral TF. Differences in handgrip strength protocols to identify sarcopenia and frailty - a systematic review. BMC Geriatr 2017;17(1):238. doi: 10.1186/s12877-017-0625-y [published Online First: 2017/10/19]

42. Charlson ME, Pompei P, Ales KL, et al. A new method of classifying prognostic comorbidity in longitudinal studies: development and validation. J Chronic Dis 1987;40(5):373-83. [published Online First: 1987/01/01]

43. Borson S, Scanlan J, Brush M, et al. The mini-cog: a cognitive ‘vital signs’ measure for dementia screening in multi-lingual elderly. Int J Geriatr Psychiatry 2000;15(11):1021- 7. [published Online First: 2000/12/13]

44. Hesterberg TMDSM, S.; Clipson, A.; Epstein, R. Bootstrap methods and permutation tests. In: A. Mdsmgpcb, ed. Intoduction to practice of statistics. New York: Palgrave Macmillan 2007.

45. Rasch D, Kubinger KD, Moder K. The two-sample t test: pre-testing its assumptions does not pay off. Statistical Papers 2011;52(1):219–31. doi: 10.1007/s00362-009-0224-x

46. Ullrich P, Eckert T, Bongartz M, et al. Life-space mobility in older persons with cognitive impairment after discharge from geriatric rehabilitation. Arch Gerontol Geriatr 2019;81:192–200. doi: 10.1016/j.archger.2018.12.007 [published Online First: 2019/01/04]

