## Supplementary material for "Validation of the German version of the Life-Space Assessment LSA-D": LSA-D Questionaire

| **DIE NÄCHSTEN FRAGEN BEZIEHEN SICH AUF IHRE AKTIVITÄTEN IN DEN LETZTEN VIER WOCHEN:** | | | **A. WIE HÄUFIG WAREN SIE IN DEN LETZTEN 4 WOCHEN IN (Name des Life-Space-Levels)?**  **Häufigkeit** | | | | **WIE SIND SIE DORT HINGEKOMMEN?** | | | | | |
| --- | --- | --- | --- | --- | --- | --- | --- | --- | --- | --- | --- | --- |
|  |  |  |  |  |  |  | **B. HABEN SIE HILFSMITTEL ODER ANDERE AUSRÜSTUNG DAFÜR VERWENDET?** | | | **C. BENÖTIGTEN SIE DAFÜR DIE HILFE EINER ANDEREN PERSON?** | | |
| WAREN SIE IN DEN LETZTEN 4 WOCHEN… | Ja | Nein | Weniger als 1 mal die Woche | 1-3 mal die Woche | 4-6 mal die Woche | Täglich | Ja | Nein | Unbekannt / keine Angabe | Ja | Nein | Unbekannt / keine Angabe |
| IN ANDEREN RÄUMEN IHRES ZUHAUSES außer dem Raum, in dem Sie schlafen?  *LIFE-SPACE 1* | **LS1** | | **LS1-Häufigkeit** | | | | **LS1-Hilfsmittel** | | | **LS1-Persönliche Hilfe** | | |
|  | ❑ | ❑ | ❑ | ❑ | ❑ | ❑ | ❑ | ❑ | ❑ | ❑ | ❑ | ❑ |
| IN DER NÄHEREN UMGEBUNG AUSSERHALB IHRER WOHNUNG (Hausflur, Terrasse, Balkon, Fahrstuhl, Hof, Garage, hauseigener Garten, Auffahrt)?  *LIFE-SPACE 2* | **LS2** | | **LS2-Häufigkeit** | | | | **LS2-Hilfsmittel** | | | **LS2-Persönliche Hilfe** | | |
|  | ❑ | ❑ | ❑ | ❑ | ❑ | ❑ | ❑ | ❑ | ❑ | ❑ | ❑ | ❑ |
| AN ORTEN IN IHRER NACHBARSCHAFT, aber außerhalb Ihrer Wohnung oder Ihrer näheren Wohnumgebung?  *LIFE-SPACE 3* | **LS3** | | **LS3-Häufigkeit** | | | | **LS3-Hilfsmittel** | | | **LS3-Persönliche Hilfe** | | |
|  | ❑ | ❑ | ❑ | ❑ | ❑ | ❑ | ❑ | ❑ | ❑ | ❑ | ❑ | ❑ |
| AN ORTEN AUSSERHALB IHRER NACHBARSCHAFT, aber innerhalb der Stadt oder der Ortschaft, in der Sie leben?  *LIFE-SPACE 4* | **LS4** | | **LS4-Häufigkeit** | | | | **LS4-Hilfsmittel** | | | **LS4-Persönliche Hilfe** | | |
|  | ❑ | ❑ | ❑ | ❑ | ❑ | ❑ | ❑ | ❑ | ❑ | ❑ | ❑ | ❑ |
| AN ORTEN AUSSERHALB DER STADT ODER DER ORTSCHAFT, IN DER SIE LEBEN?  *LIFE-SPACE 5* | **LS5** | | **LS5-Häufigkeit** | | | | **LS5-Hilfsmittel** | | | **LS5-Persönliche Hilfe** | | |
|  | ❑ | ❑ | ❑ | ❑ | ❑ | ❑ | ❑ | ❑ | ❑ | ❑ | ❑ | ❑ |
